# The physiological and clinical-behavioral effects of HRV biofeedback in adolescents with autism: a pilot randomized controlled trial

**DOI:** 10.1101/2023.05.31.23290775

**Authors:** Anoushka Thoen, Kaat Alaerts, Jellina Prinsen, Jean Steyaert, Tine Van Damme

**Author notes:** Correspondence to Anoushka Thoen, KU Leuven, Department of Rehabilitation Sciences, Research Group for Adapted Physical Activity and Psychomotor Rehabilitation, Herestraat 49-mailbox 1510, 3000 Leuven, Belgium.

## Abstract

**Background:** Adolescents with autism present lower levels of cardiac vagal modulation in comparison to typically developing peers. These lower values are also associated with psychosocial and behavioral problems. Heart Rate Variability Biofeedback (HRVB) was therefore suggested as an interesting avenue for further exploration since it focusses on the up-regulation of cardiac vagal modulation and has demonstrated positive effects on mental health outcomes. However, scarce evidence was present regarding the effectiveness of HRVB in this population. It was hypothesized that HRVB would increase the level of cardiac vagal modulation in adolescents with autism which would result in positive effects on physiological outcome measures and psychosocial parameters. Furthermore, it was hypothesized that a home-based, non-supervised HRVB training would be feasible in this population.

**Methods:** A single-blind, randomized sham-controlled pilot trial was used. During the initial single-blind phase, adolescents with autism performed supervised HRVB (n=24) or a sham training (n=20) for five weeks with one guided session per week and home-based practice (20 min) on the remaining days. In the subsequent follow-up period of five weeks, half of the adolescents of each group received HRVB training at home, in a non-supervised manner, whereas the other subset did not practice during that period. A combination of physiological, cortisol and behavioral data was collected during standardized stress-provoking assessments before (T0) and after each training period (T1 and T2).

**Results:** Supervised HRVB resulted in a late increase in cardiac vagal modulation in adolescents with autism. Heart rate increased and cortisol decreased significantly immediately after the supervised HRVB training, but none of these effects remained after the follow-up period of five weeks. None of the outcome measures on psychosocial functioning and self-reported stress revealed a significant change following the supervised HRVB training. The home-based HRVB training was feasible in this population but with a significant decrease in compliance rate. However, adolescents reported significantly lower symptoms of stress after this training period, regardless of the prior training (sham or HRVB).

**Conclusion:** HRVB is feasible and effective in adolescents with autism as demonstrated by late-emerging increases in cardiac vagal modulation and less self-reported symptoms of stress. Researchers are encouraged to replicate this study with a larger sample size and to further explore the possible working mechanisms of HRVB.

## Introduction

Autism Spectrum Disorder (ASD)^1^ is a neurodevelopmental condition characterized by persistent deficits in social communication and interaction as well as the presence of restricted, repetitive behaviors and sensory anomalies (American Psychiatric Association, 2022). The prevalence in developed countries is estimated to be at least 1.5% (Lyall et al., 2017). Moreover, individuals with autism report more co-occurring mental health problems than the general population (Lai et al., 2019). For instance, pooled prevalence rates from a recent meta-analysis indicated a prevalence of 20% for anxiety disorders, 11% for depressive disorders and 0-3.6% for trauma and stressor-related disorders across the entire lifespan (Lai et al., 2019). In the adolescent period, individuals with autism are even more vulnerable for stress-related difficulties and mental health problems, considering that this period is typically characterized by rapid changes in several developmental domains (e.g.: social context, affect regulation and behavior; Picci & Scherf, 2015). This implies that mental health assessment and treatment should be an integral part of the clinical care in this population (Lai et al., 2019). To date, multiple studies have shown improved mental health outcomes after mind-body interventions such as yoga, mindfulness and acceptance and commitment therapy in individuals with autism (Hartley et al., 2019; Hourston & Atchley, 2017; Semple, 2019). However, amongst others, one of the reported problems regarding these interventions is the need for modification of instructions as many mind–body interventions frequently use figurative and abstract language in addition to relying on a certain amount of imagination skills. This can be challenging for individuals with autism, who are more likely to take language literally. In addition, most of these interventions rely on interoceptive abilities, which can be underdeveloped in certain individuals with autism and may therefore result in lower treatment efficacy (Dubois, 2016; Williams et al., 2022).

Therefore, it has been suggested that interventions based on breathing techniques such as Heart Rate Variability biofeedback (HRVB) might be relevant and beneficial for adolescents with autism (Condy et al., 2019; Coulter et al., 2022; Heiss et al., 2021; Patriquin et al., 2019). In short, the training aims at maximally increasing Heart Rate Variability (HRV), by slowing down the breathing frequency, while providing continuous feedback on HRV. Increasing HRV can be achieved by breathing on an individual’s resonance frequency, which refers to the breathing frequency at which breathing-related influences and baroreflex-related influences on heart rate resonate, thereby maximally increasing HRV (Lehrer & Gevirtz, 2014). Since both influences are vagally (or parasympathetically) mediated, an increase in HRV reflects increased cardiac vagal modulation (Laborde et al., 2022; Sevoz-Couche & Laborde, 2022). Ultimately, during HRVB, the baroreflex is strengthened which results in cardiovascular benefits while the increased cardiac vagal modulation is suggested to underlie psychological and cognitive benefits (Lehrer et al., 2020; Lehrer & Gevirtz, 2014; Sevoz-Couche & Laborde, 2022). For a detailed overview of possible working mechanisms, readers are referred to review articles of Lehrer and Gevirtz (2014) and Sevoz-Couche and Laborde (2022).

In children and adolescents with autism, the overall levels of cardiac vagal modulation are lower in comparison to typically developing peers (Cheng et al., 2020; Makris et al., 2022; Thoen et al., 2023). In addition, lower levels of cardiac vagal modulation are associated with higher severity of autism characteristics (Arora et al., 2021; Condy et al., 2019), the presence of internalizing and externalizing behavior (Arora et al., 2021; Chiu et al., 2023), lower social-emotional skills (Arora et al., 2021; Makris et al., 2022; Patriquin et al., 2019) and the presence of more severe auditory and visual hyperreactivity (Matsushima et al., 2016). Consequently, a training that specifically targets the up-regulation of cardiac vagal modulation, such as HRVB (Lehrer & Gevirtz, 2014), is an interesting avenue for further exploration (Coulter et al., 2022; Matsushima et al., 2016; Neuhaus et al., 2016; Patriquin et al., 2014). Furthermore, a dysfunction of the Hypothalamic-Pituitary-Adrenal axis (HPA) – a neuro-endocrine system involved in the stress response with cortisol as its main effector – has also been demonstrated in this population (Makris et al., 2022). Given the anatomical link between the autonomic nervous system and the HPA axis (Thayer & Lane, 2000), it could be assumed that the hypothesized up-regulation of cardiac vagal modulation may also result in changes of the HPA axis functioning.

Previous research indicates that HRVB can reduce symptoms of anxiety and depression in both clinical and non-clinical populations (Goessl et al., 2017; Lehrer et al., 2020; Lehrer & Gevirtz, 2014). In comparison to the previously mentioned mind-body interventions, HRVB is digitalized and not associated with figurative language, nor does it rely on well-developed interoceptive abilities. In contrast, the structured and visual set-up of this training may facilitate the communication and comprehension of the instructions for adolescents with autism. HRVB also has the potential to be transformed into a home-based training using applications on mobile devices. This might be beneficial for this particular population, as it would allow the adolescents to practice in their familiar environment and to facilitate the integration and transfer into their daily routine (Bölte et al., 2010; Lamash et al., 2023). Recently, the feasibility of HRVB in children and adolescents with autism has been reported (Coulter et al., 2022; Goodman et al., 2018). In the study of Goodman et al. (2018), children and adolescents (10-15 years) received either HRVB (n=7) or combined HRVB with neurofeedback (n=8). In total, 12 hours of lab sessions were spread over the course of 6 weeks (1×2hours/week or 2×1hour/week), supplemented with home practice for 10-20 minutes per day. Within-group analyses for the HRVB group showed improvements in emotion regulation and social behavior. However, no significant between-group differences were found in addition to non-significant improvements on measures of cardiac vagal modulation after the HRVB training. Findings from a recent pilot randomized controlled trial on home-based HRVB during 12 weeks (2 guided sessions of 30 minutes at the start) in adolescents and young adults with autism (n=15; 13-22 years) revealed a significant reduction of self-reported anxiety (Coulter et al., 2022). Both studies used small sample sizes, varying training protocols and outcome measures in addition to only one study that included a control group. These differences make it difficult to generalize the results for individuals with autism and hinder comparison with research findings in other populations.

Therefore, to further explore the efficacy of HRVB in adolescents with autism and to examine potential effects on psychosocial outcomes and physiological parameters, a single-blind, randomized sham-controlled pilot trial, with between-subject design was used. During the initial single-blind phase, adolescents with autism performed supervised HRVB or sham training for five weeks with one supervised session per week and home-based practice on the remaining days. In the five weeks after the supervised training, half of the adolescents of each group (HRVB and sham) performed HRVB at home, in a non-supervised manner, whereas the other subset will not perform any training (See Figure 1). The primary hypotheses were defined as follows: (1) supervised HRVB enhances cardiac vagal modulation in adolescents with autism; (2) supervised HRVB results in positive changes in behavior, mental health outcomes and/or physiological changes (heart rate, breathing frequency and cortisol levels); (3) home-based, non-supervised HRVB is feasible in adolescents with autism.

**Fig. 1.**
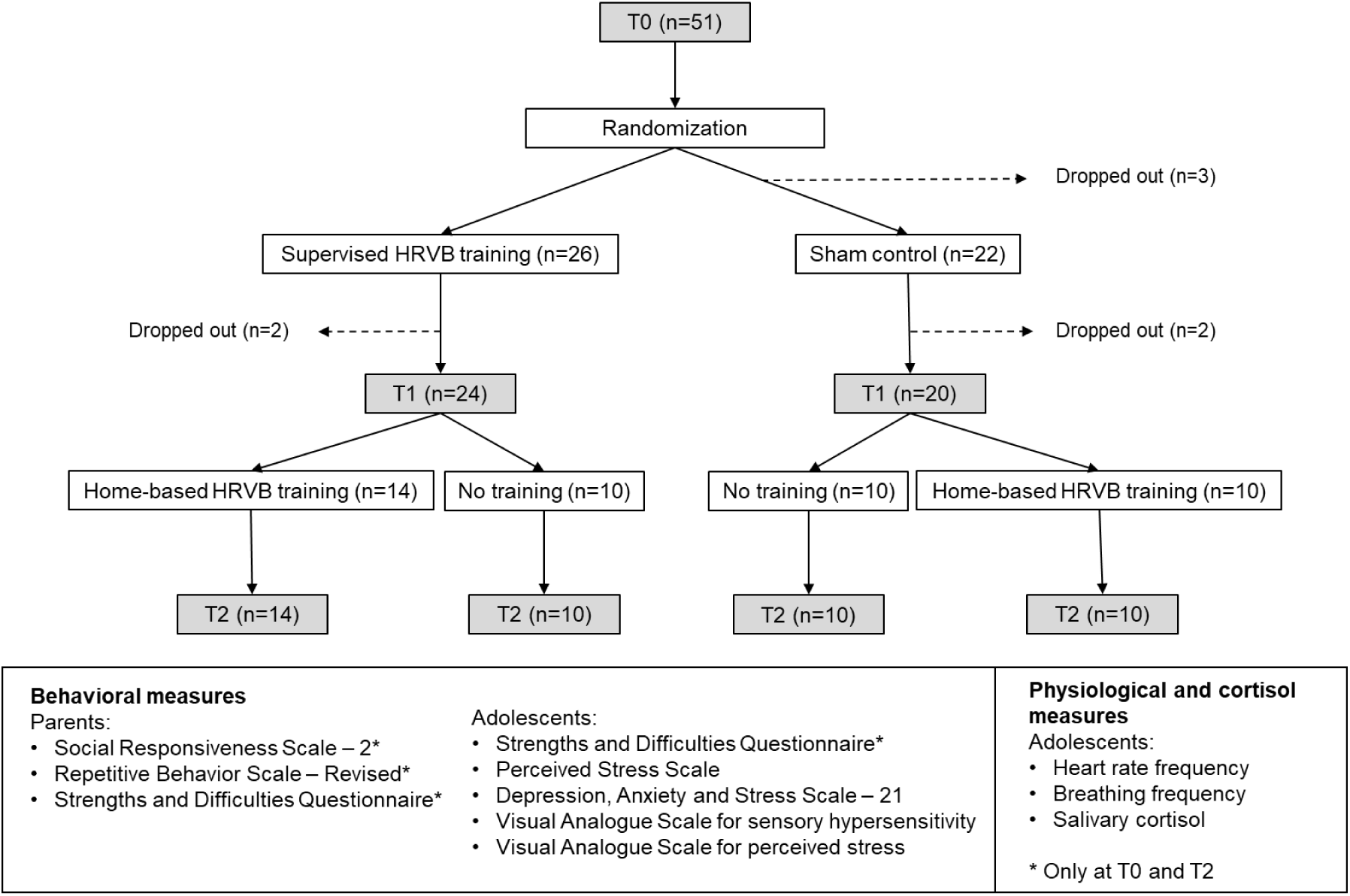
Flowchart of the Study. Note: HRVB=Heart Rate Variability Biofeedback.

## Methods

### General procedure

A pilot randomized-controlled trial design was used for the supervised and home-based, non-supervised HRVB training (See Figure 1). All adolescents performed a baseline measurement (Time 0 (T0)) before being randomly assigned to the supervised HRVB group or the sham-control group. After five weeks of training, all adolescents were re-assessed (T1) and afterwards, re-assigned to receive five weeks of home-based, non-supervised HRVB training (HT) or no training (NT). After these five weeks, a follow-up assessment (T2) was performed. Block procedures were used for randomization and groups were balanced on sex and severity of autism characteristics (based on SRS-2 total t-scores in a Dutch-speaking autism population; Constantino & Gruber, 2015).

### Ethical approval and informed consent

This study was performed in line with the principles of the Declaration of Helsinki and was approved by the Ethics Committee UPC KU Leuven (ref. EC2020-541) and the Ethics Committee Research UZ/KU Leuven (ref. S64219). Informed parental consent and adolescent assent was obtained before the first study visit. The data was collected as part of a larger study as described in Thoen et al. (2021) and has been registered in the online clinical study repository ClinicalTrials.gov (ID: NCT04628715).

### Participants

Adolescents aged between 13 and 18 years were recruited from December 2019 until September 2022 across Flanders (Belgium), facilitated by the Leuven Autism Expertise Centre and the Leuven Autism Research Group (KU Leuven). In addition, (special education) schools, autism advocacy organizations and support groups as well as independent clinical practices were approached to support recruitment. A formal diagnosis of ASD was required, based on the criteria of the Diagnostic and Statistical Manual of Mental Disorders IV-TR/5 (DSM-IV-TR/5; American Psychiatric Association, 2000, 2013). The use of medication and the presence of co-occurring disorders was registered. Exclusion criteria were (i) the presence of an intellectual disability as described in the DSM-IV-TR/5; (ii) the presence of an uncorrected hearing- or vision impairment; (iii) the presence of congenital heart diseases, diagnosed cardiovascular abnormalities or somatic diseases with a known impact on heart function; (iv) the presence of acute agitation and/or acutely impairing psychiatric symptoms (psychosis, mania or major depressive disorder); (v) pregnancy; (vi) insufficient knowledge of the Dutch language and (vii) participation in other clinical trials.

Following the recommendations of Whitehead et al. (2016) for a pilot RCT with medium effect size (α=0.05; 1-β=0.80), a priori sample size calculations resulted in 40 adolescents to be recruited i.e. to obtain 10 adolescents receiving either supervised HRVB or sham with or without additional home-based training (HRVB+HT; HRVB+NT; Sham+HT; Sham+NT). The expected dropout rate was 20%, thereby increasing the sample size up to at least 48 adolescents.

### Assessments

Each assessment session (T0, T1, T2) lasted one hour and included a preparatory phase to attach the sensors for physiological data-collection and measurements of physiology during a 30-minute stress-provoking protocol. At regular time points during the assessment, salivary samples were collected for determination of cortisol levels. A detailed description of the assessment (f.i. standardized body positioning and outcome measures) has been described in Thoen et al. (2021).

In short, during the baseline period (the first ten minutes of the stress-provoking protocol), the adolescents had to sit quietly with their eyes closed. The first five minutes were excluded from data-analyses to incorporate a corresponding acclimatization period as recommended by Quintana et al. (2016). Subsequently, the ‘Stroop Word-Color Interference task’ (Kushki et al., 2013) and the ‘Social Stress Recall Task’ (SSRT; Bishop-Fitzpatrick et al., 2017; Richman et al., 2007) were used as stress-provoking tasks, each followed by a 5-minute resting period. Both tasks had a duration of five minutes and have been used previously to provoke stress in individuals with autism (Bishop-Fitzpatrick et al., 2017; Kushki et al., 2013). Accordingly, physiological data were obtained during five 5-min recording conditions in the following sequential order: ‘baseline’; ‘Stroop task; ‘rest 1’, ‘SSRT’ and ‘rest 2’.

#### Physiological measurements

All physiological data were captured using the NeXus-10 MKII biofeedback device and Biotrace+ Software (Mind Media B.V., The Netherlands). Three-lead electrocardiographic data were collected with disposable, self-adhesive and pre-gelled electrodes (Kendall™ ECG Electrodes Arbo™ H124SG, Covidien, Ireland) without specific skin preparations at a sampling rate of 256 SPS. The negative electrode was placed medial to the right coracoid process, just below the clavicle. The neutral electrode was placed similarly on the contralateral side. The positive electrode was placed at the lower left ribcage. For the recording of the breathing frequency, an elastic band with stretch-sensitive sensors and a sampling rate of 32 SPS was placed around the waist.

#### Cortisol salivary sampling

Salivette® Cortisol cotton swabs (Sarstedt Inc., Rommelsdorf, Germany) were used for the collection of saliva samples at three time points to determine the level of cortisol. To rule out the influence of the cortisol awakening response, the assessments were always performed in the afternoon (Hollocks et al., 2014). In parallel with the physiological data recordings, a baseline salivary sample was collected before the start of the stress-provoking protocol. Next, the second and third saliva samples were collected at 20 minutes after each of the stress-provoking tasks (Corbett et al., 2019). This time window was chosen since salivatory cortisol levels are known to peak at around 10-30 minutes after stress cessation (Foley & Kirschbaum, 2010). Upon collection, samples were stored under appropriate controlled conditions (−20°C) and analyzed with the commercial enzyme immunoassay Cortisol ELISA kit of Salimetrics, Europe.

#### Psychosocial functioning and perceived stress

The parent-reported Social Responsiveness Scale (SRS-2; Constantino & Gruber, 2015; Constantino & Gruber, 2012) and Repetitive Behavior Scale (RBS-R Dutch translation; Lam & Aman, 2007) were used to assess autism characteristics. Furthermore, psychosocial problems and strengths were assessed using the parent- and self-reported Strengths and Difficulties Questionnaire (SDQ Dutch translation; Goodman, 2001). Self-report tools were filled out by the adolescents to report on perceived stress levels over the last month (Perceived Stress Scale – Adolescent version; Van der Ploeg, 2013) and the presence of stress symptoms over the last week (stress subscale from the Depression, Anxiety and Stress Scale – 21; de Beurs et al., 2001). The level of sensory hypersensitivity was assessed using a visual analogue scale going from zero (no sensory hypersensitivity) to ten (severe sensory hypersensitivity and negative impact on functioning). The level of perceived stress for each of the stress-provoking tasks was also evaluated using a visual analogue scale ranging from zero (no stress) to ten (highest level of perceived stress possible). All questionnaires were filled out online, maximally 1 week prior to the assessments, except for the visual analogue scale on perceived stress, which was administered after each assessment session (See Figure 1).

### Training

#### Supervised HRVB training

The training was based on the biofeedback protocol provided by Lehrer and colleagues (2013). The supervised HRVB group followed a fixed scheme of one supervised session per week (30 minutes) and home-practice on the other days (20 minutes). During the first training session, the personal resonance frequency of the adolescents was determined using a standardized protocol designed by Mind Media B.V. and Richard Gevirtz (Lehrer et al., 2013; Lehrer et al., 2000; Shaffer & Meehan, 2020). This frequency refers to a breathing frequency between 4.5 and 7 breaths per minute at which heart rate and the breathing pattern are in phase, also known as Respiratory Sinus Arrhythmia (RSA; Lehrer et al., 2020). All adolescents were instructed to breathe at their individually determined resonance frequency through stepwise guidance from the researcher using appropriate breathing techniques (pursed lips and abdominal breathing). During the sessions, the adolescent’s heart rate and breathing frequency were visualized on a computer screen in addition to the breathing pacer that they had to follow. From the third session onwards, the adolescents were instructed to ignore the breathing pacer and to breathe in phase with their heart rate. However, they did continue breathing at their resonance frequency during home practice for five weeks using a breathing pacer application (Awesome Labs LLC, 2020; Trex, 2015). They were asked to rate the level of comfort and compliance daily during home-practice via a secure web-based application (m-path, KU Leuven).

The sham-control group followed the same scheme as the supervised HRVB group (5 weeks). However, the adolescents practiced at a normal breathing frequency (between 9 and 16.2 breaths per minute) during the supervised sessions as well as during home-practice, thus faster than their resonance frequency. The resonance frequency was first determined, followed by gradually increasing the pacer’s frequency until a faster but comfortable breathing frequency was obtained.

#### Home-based HRVB training

The five weeks of home-based, non-supervised HRVB started with one visit at T1. During this visit, the adolescent’s resonance frequency was verified and adapted accordingly, when needed. Next, the adolescents practiced daily at their resonance frequency for 20 minutes using a biofeedback smartphone application (©Elite HRV, Asheville, North Carolina, USA). In addition, a compatible heart rate sensor (Polar® H10, Polar, Finland) monitored the adolescent’s heart rate. In case the adolescents experienced problems with this heart rate sensor and/or the biofeedback smartphone application, they were instructed to breathe at their resonance frequency by using the breathing pacer from the supervised HRVB training. Finally, a secure web-based application (m-path, KU Leuven) was used to rate their level of comfort and compliance on a daily basis.

The control group was instructed not to practice during these five weeks.

### Data preprocessing

Heart rate data were preprocessed using Kubios HRV Premium (version 3.4.3, University of Eastern Finland, Kuopio, Finland) using the automatic artifact correction algorithm and medium automatic noise detection. In addition, visual inspection of the raw data was performed. Analysis of the data regarding HRV was also performed with Kubios HRV Premium using RSA as a measure of cardiac vagal modulation. This was calculated using the time domain measure ‘Root Mean Square of Successive Differences between normal heartbeats’ (RMSSD) and the frequency domain measure ‘High Frequency Heart Rate Variability’ (HF-HRV; Task Force, 1996; Laborde et al., 2017). Researcher-developed scripts in MATLAB R2020b (MathWorks, Natick, Massachusetts, USA) were used for preprocessing and calculation of the mean respiratory frequency for the baseline period in the stress-provoking protocol during each assessment (T0, T1, T2).

### Analyses

Statistical analyses were conducted using SPSS software (IBM SPSS Statistics, version 27) and Statistica (Tibco software Inc. version 14). Physiological and cortisol data were logarithm-transformed when needed to meet normality assumptions prior to data analyses.

Differences in sociodemographic, psychosocial functioning and self-perceived stress between the groups were examined using: (1) independent sample t-tests or Mann-Whitney U-tests and chi-square tests of independence when comparing two groups at T0 (HRVB versus sham) and (2) one-way ANOVA (with Tukey’s HSD as post-hoc analysis) or Kruskal-Wallis H tests and chi-square tests of independence when comparing four groups at T1 (HRVB+HT, HRVB+NT, Sham+HT and Sham+NT).

A mixed-effect analysis of variances was used to examine the effect of the supervised HRVB training on cardiac vagal modulation (separate models for RMSSD and HF-HRV). The factor ‘subject’ was used as a random effect, whereas the factors ‘supervised training group’ (HRVB, sham), ‘assessment session’ (T1, T2), and ‘condition’ (baseline, Stroop, rest1, SSRT, rest2) were inserted as fixed effects. To correct for variance in the adolescents’ T0 scores, these pre-training values were included as a covariate in the model. Post-hoc analyses were Bonferroni-corrected. Next, to examine the potential additional effect of the home-based, non-supervised training on cardiac vagal modulation, directly on the T2 outcome, mixed-effects analyses were conducted on the T2 outcome data with the factor ‘subject’ as random effect, and the factors ‘supervised training group’ (HRVB, sham), ‘home training’ (yes, no), and ‘condition’ (base, Stroop, rest1, SSRT, rest2) as fixed effects.

The same mixed-effect analyses were performed on heart rate frequency and breathing frequency data following the supervised and home-based HRVB training. In addition, for the cortisol data, similar mixed-effect analyses were performed, i.e., with the random factor ‘subject’, and the fixed factors ‘supervised training group’ (HRVB, sham), ‘assessment session’ (T1, T2), and ‘condition’ (post-baseline, post-Stroop, post-SSRT) and T0 data as a covariate. To examine the potential additional effect of the home-based, non-supervised training on cortisol, directly on the T2 outcome, the additional fixed factor ‘home training’ (yes, no) was added to the model.

Mixed-effect analyses were also performed for the total scores of the questionnaire data assessing psychosocial functioning and stress using similar models as mentioned above but without the fixed factor ‘condition’. Since for the SRS, RBS and SDQ, outcomes were only assessed at T0 and T2, a model was constructed using only the T2 data (with T0 data as a covariate).

For completeness, exploratory within-group analyses were performed for each group separately using dependent sample t-tests or Wilcoxon Signed-Rank Test on pre- to post-difference total scores for each of the outcome measures on psychosocial functioning and self-perceived stress as well as for heart rate, cardiac vagal modulation (RMSSD and HF-HRV), breathing frequency and cortisol level during each assessment (T0, T1 and T2). Cohen’s d and r were used as measures of effect size, respectively. Hedges’ correction was applied on Cohen’s d in case the sample size was smaller than 20.

To examine the compliance rate between the supervised HRVB group versus the sham group between T0 and T1, a Mann-Whitney U-test was used. To examine the compliance rate between the supervised (T0-T1) and home-based HRVB training (T1-T2), the Wilcoxon Signed-Rank Test was performed with the Sham+HT and HRVB+HT groups only. Compliance rate was defined as the number of minutes spent on home-practice divided by the maximum amount of minutes that the adolescents could practice during their training period, represented as a percentage.

Missing values on one or more outcome measures were excluded analysis per analysis.

## Results

The randomization of the adolescents across the groups, number of dropouts and final number of adolescents in each training group is visualized in Figure 1. At T2, the physiological and cortisol data collection was discontinued in one adolescent due to mental health problems and were therefore treated as missing data. Also, at T2, for one adolescent, the cortisol values were too low to be detected by the analysis kit and therefore defined as missing data. Demographic and diagnostic information per training group is presented in Table 1. At T0, there were no significant group differences for age, mean levels of physiological parameters and cortisol or any of the outcomes of the self- or parent reported questionnaires (*p* >.05). Use of the following medications was reported at T0: contraceptive medication (n=5), sleep medication (n=1), medication related to allergies, somatic or neurological problems (n=8), methylphenidate (n=5), non-tricyclic antidepressants (n=9), antipsychotic medication (n=10) and anti-epileptic medication (n=3). During the intervention, two adolescents started taking methylphenidate; one adolescent’s use of methylphenidate was discontinued as well as another adolescent’s use of antipsychotic medication. Co-occurring neurodevelopmental and/or psychiatric disorders were present in 19 adolescents, consisting of: attention deficit (hyperactivity) disorder (n=9), obsessive-compulsive disorder (n=3), bipolar disorder (n=1), depression (n=4), anxiety disorder (n=4), eating disorder (n=1), trauma (n=1), learning disorder (n=1), Tourette Syndrome (n=1) and developmental coordination disorder (n=3).

**Table 1.** Baseline characteristics of adolescents with autism randomized to receive the supervised HRVB or sham training.

|  | HRVB<br>(n=26; 14 females) |  |  | Sham<br>(n=25; 16 females) |  |  | *p* |
| --- | --- | --- | --- | --- | --- | --- | --- |
|  | Mean | SD | Range | Mean | SD | Range |  |
| Age (years) | 15.64 | 1.63 | 12.5-18.1 | 16.12 | 1.73 | 13.3-18.5 | .309<sup>a</sup> |
| SRS-2 total | 85.58 | 14.35 | 58-111 | 89.76 | 15.26 | 56-111 | .318 |
| RBS-R total | 16.85 | 10.53 | 1-41 | 22.12 | 13.85 | 2-52 | .131 |
| SDQ-P total | 16.12 | 6.79 | 4-30 | 15.64 | 4.93 | 8-24 | .533<sup>a</sup> |
| SDQ-S total | 16.12 | 5.81 | 4-26 | 16.68 | 6.29 | 6-28 | .740 |
| DASS stress subscale | 10.62 | 4.63 | 3-21 | 9.32 | 4.77 | 0-21 | .330 |
| PSS | 23.62 | 7.06 | 8-38 | 24.44 | 6.46 | 13-34 | .666 |
| Sensory hypersensitivity | 6.31 | 2.67 | 1-10 | 5.84 | 2.73 | 1-10 | .537<sup>a</sup> |
| RMSSD base (ms) | 44.92 | 34.80 | 5-170 | 34.98 | 16.82 | 11-86 | .486<sup>a</sup> |
| Heart rate base (bpm) | 84.22 | 12.79 | 63-118 | 83.44 | 8.78 | 66-101 | .801 |
| Breathing frequency base (/min) | 16.65 | 2.95 | 10-22 | 18.00 | 3.29 | 11-24 | .130 |
| Cortisol base (µg/dl) | 0.11<sup>b</sup> | 0.06 | 0.04-0.27 | 0.17 | 0.13 | 0.03-0.56 | .426 |

a Non-parametric Mann-Whitney U-Test was used; b Missing value (n=1). Note: base=baseline; DASS=Depression Anxiety and Stress Scale; HRVB=Heart-Rate Variability Biofeedback; PSS=Perceived Stress Scale; RBS-R=Repetitive Behavior Scale–Revised; RMSSD=root mean square of successive differences; SD=Standard Deviation; SDQ=Strengths and Difficulties Questionnaire; SRS-2=Social Responsiveness Scale–Second Edition

During the supervised training period, the actual number of home-training days deviated from the foreseen 31 days to a range of 29 to 33 days (mean 30 days; SD 0.77). Training compliance did not differ significantly between the HRVB group (69±26%, range 7-100%) and the sham group (78±18%, range 26-97%; *U*=208, *p*=.450). The resonance frequency for the HRVB group ranged between 5.5 and 9 breaths per minute (mean 6.75; SD 0.79) while the sham HRVB group practiced at a breathing frequency ranging between 9.2 and 16.2 breaths per minute (mean 10.90; SD 1.77).

During the following five-week period of home-based training, the actual amount of maximum practice days deviated from the foreseen 35 days to a range of 32 to 35 days (mean 34 days; SD 1.17). Training compliance did not differ significantly between the two groups that performed the home-based training (HRVB+HT=41±38% (range 1-99%) and Sham+HT=67±22% (range 37-97%); *U*=43, *p*=0.122). During this training period, the resonance frequency for the adolescents ranged between 5.5 and 8 breaths per minute (mean 6.79; SD 0.59).

There was a significant decrease in the compliance rate from the supervised HRVB and sham training between T0 and T1 (76±20%) to the home-based HRVB training between T1 and T2 (52±34%; Z=-3.743, *p*<.001, *r*=0.764).

### Effect of HRVB training

#### Primary outcome – cardiac vagal modulation

The mixed-effects analyses of variances examining the effect of HRVB versus sham on RMSSD revealed a significant ‘supervised training group by assessment session’ interaction effect (*F*(1,372)=10.880, *p*=.001, η^2^_p_ =0.026). This indicated significantly higher RMSSD in the HRVB training group, compared to the sham group, but only at the follow-up session (T2; *p_Bonferroni_*<.001), not immediately after the training (T1; *p*=1.00). No main or interaction effect of ‘condition’ was revealed (all, *p*>.05), indicating that the observed effect of HRVB training at T2 was evident across conditions (Figure 2A visualizes the effect across conditions, Supplementary Figure 1A visualizes the effect separately for each condition, i.e., baseline, Stroop task, rest1, SSRT, rest2). None of the other main and interaction effects were significant (all, *p*>.05). A similar pattern of results was revealed for the HF-HRV assessment of cardiac vagal modulation (see Supplementary Figure 2).

**Fig. 2.**
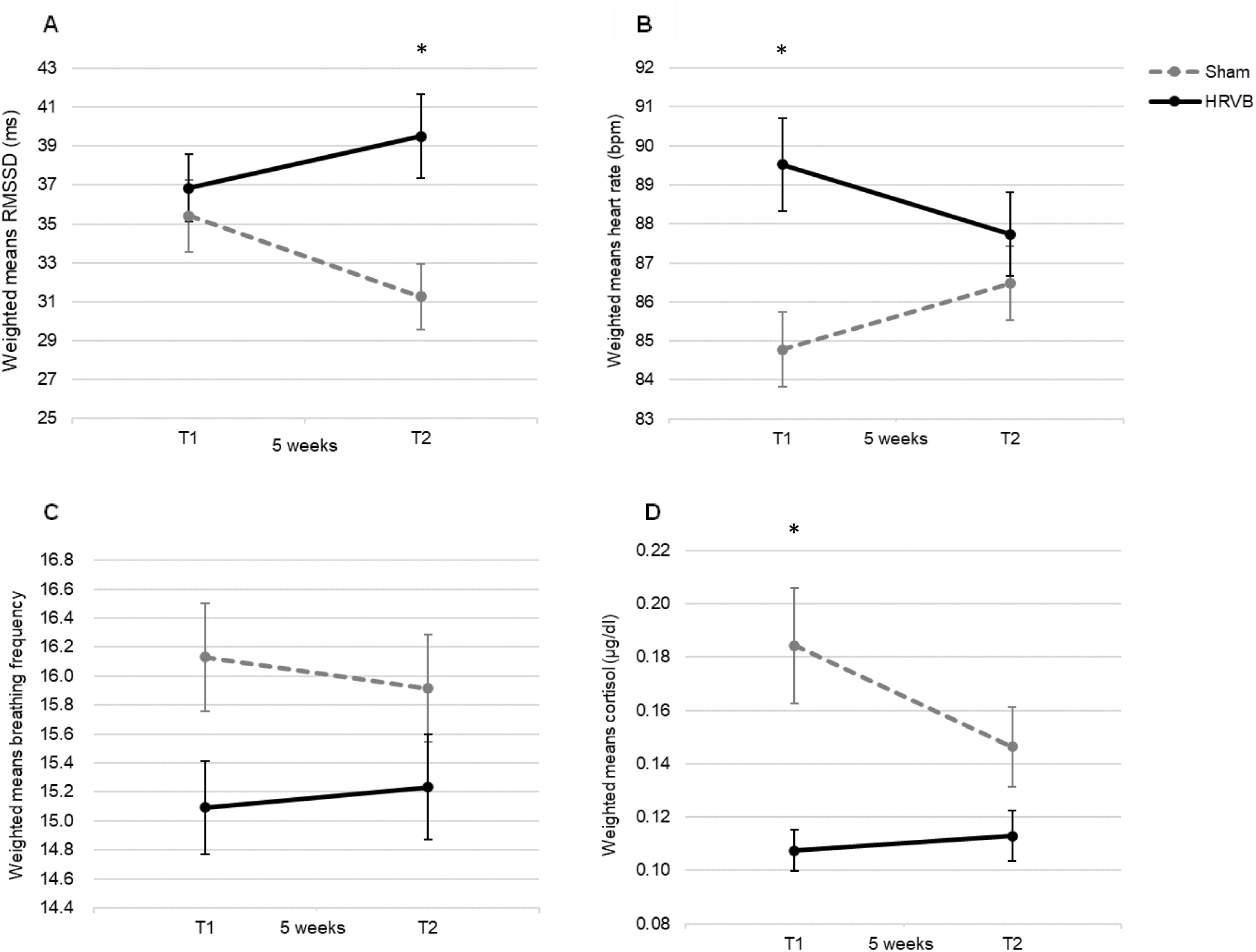
Effect of HRVB training on physiological outcome measures and cortisol levels. Weighted means are visualized separately for the HRVB and sham training group at assessment sessions T1 and T2 (* p<.001). Panel A: Effect on cardiac vagal modulation, assessed using RMSSD. Panel B: Effect on heart rate. Panel C: Effect on breathing frequency. Panel D: Effect on cortisol level. Note: bpm=beats per minute; HRVB=Heart Rate Variability Biofeedback; RMSSD=Root Mean Square of Successive Differences

Next, mixed-effect analyses were performed, examining the potential additional effect of the home-based training on cardiac vagal modulation (at the T2 outcome). This analysis revealed no significant main effect of ‘home-training’ (*F*(1,155)=0.297, *p*=.589, η^2^_p_ =0.008), nor any significant interaction with this factor (all, *p* >.05). This indicates that the heightened RMSSD at T2 observed in the HRVB training group was uniform, irrespective of the additional five-week home-training (provided between T1 and T2).

#### Secondary outcome – heart rate frequency and breathing frequency

A significant ‘supervised training group by assessment session’ interaction effect was found on heart rate frequency (*F*(1,372)=10.728, *p*=.001, η^2^_p_=0.026), indicating a significantly higher heart rate in the HRVB training group compared to the sham group but only at T1 (*p_Bonferroni_*<.001), not at the follow-up session (T2, *p_Bonferron_*_i_=.427). No main or interaction effect of ‘condition’ was revealed (all, *p*>.05), indicating that the observed effect was evident across conditions (Figure 2B visualizes the effect across conditions, Supplementary Figure 1B visualizes the effect separately for each condition, i.e., baseline, Stroop task, rest1, SSRT, rest2). In contrast, a main effect of ‘condition’ was found on breathing frequency (*F*(4,372)=11.954, *p*<.001, η^2^_p_=0.104) indicating a significantly higher breathing frequency during the Stroop task with respect to the other conditions (*p_Bonferroni_*<.001) and a significantly higher breathing frequency during the baseline condition with respect to rest1 and rest2 (*p_Bonferroni_*<0.05). None of the other main or interaction effects were significant on breathing frequency (all, *p*>.05; Figure 2C visualizes the effect across conditions, Supplementary Figure 1C visualizes the effect separately for each condition, i.e., baseline, Stroop task, rest1, SSRT, rest2).

Mixed-effect analyses, examining the potential additional effect of the home-based training on the T2 outcome assessment, did not reveal any main or interaction effects with the factor ‘home-training’, indicating no significant further modulation of heart rate frequency and breathing frequency by this additional training (all, *p*>.05).

#### Secondary outcome – salivary cortisol

Mixed-effects analyses of the cortisol salivary data revealed a significant ‘training group by assessment session’ interaction effect (*F*(1,199)=7.644, *p*=.006, η^2^_p_=0.034), indicating significantly lower cortisol levels in the HRVB training group, compared to the sham group, but only significant at T1, immediately after the training (*p_Bonferroni_*<.001). At T2, the effect was trend significant (*p_Bonferroni_*=.06, see Figure 2D). Similar to the RMSSD data, no significant main or interaction effect with ‘condition’ was revealed (all, *p*>.05), indicating that the aforementioned effect on cortisol levels was evident irrespective of the condition (Figure 2D visualizes the effect across conditions, Supplementary Figure 1D visualizes the effect separately for each condition, i.e., post-baseline, post-Stroop task, post-SSRT).

Mixed-effect analyses, examining the potential additional effect of the home-based training on the T2 outcome assessment, did not reveal any main or interaction effects with the factor ‘home-training’, indicating no significant further modulation of cortisol levels by this additional training (*p*>.05).

#### Secondary outcomes – questionnaires

Mixed-effect analyses of the DASS stress subscale and PSS, assessed at T0, T1 and T2 did not reveal a main effect of ‘supervised training group’ ((*F*(1,42)=0.245, *p*=.624, η^2^_p_=0.006); (*F*(1,42)=1.225, *p*=.275, η^2^_p_=0.029), resp.) nor significant interaction effects with assessment session (all, *p*>.05), indicating no significant modulation of self-reported psychological stress upon the HRVB training versus the sham training. A significant ‘supervised training group by assessment session’ interaction effect was found for the VAS of the Stroop task (*F*(1,40)=7.351, *p*=.009, η^2^_p_=0.142), but post-hoc analyses were non-significant (*p_Bonferroni_*>.05). No such effect was found for the VAS of the SSRT (*F*(1,40)=0.612, *p*=.439, η^2^_p_=0.014). None of the other main and interaction effects were significant for both VAS scores (all, *p*>.05). For sensory hypersensitivity, a significant main effect of ‘session’ was found (*F*(1,42)=5.113, *p*=.029, η^2^_p_=0.109), indicating lower self-reported levels of sensory hypersensitivity at T2 as opposed to T1. There were, however, no significant other main effects or interaction effects meaning that lower scores were present in both groups, irrespective of the received training (*p*>.05).

For the questionnaires assessed only at T0 and T2 (SRS, RBS, SDQ), no significant main effects with the factor ‘supervised training group’ were revealed, indicating no significant effect of the training on these scales examining autism characteristics and behavioral symptoms (main effect SRS (*F*(1,39)=0.482, *p*=.492, η^2^_p_=0.012); RBS (*F*(1,39)=0.805, *p*=.554, η^2^_p_=0.485); SDQ-Parent (*F*(1,39)=0.052, *p*=.821, η^2^_p_=0.001); SDQ-Self (*F*(1,39)=1.488, *p*=.230, η^2^_p_=0.037). None of the other main and interaction effects were significant (all, *p*>.05). Mixed-effect analyses, examining a potential additional effect of the home-based training at the T2 outcome did not reveal any significant modulation for most of the questionnaires (all, *p*>.05), except for the DASS stress subscale (*F*(1,0)=4.298, *p*=.045, η^2^_p_=0.102), indicating lower self-reported stress at T2 for adolescents in the home-training group with respect to adolescents who received no home-training. None of the other main and interaction effects were significant (all, *p*>.05).

### Exploratory within-group differences

Tables 2–4 report all mean raw scores and exploratory within-group differences on physiological, cortisol and questionnaire data (Supplement 3 provides exploratory within-group differences on physiological and cortisol data per condition, per session). For physiological parameters (means across conditions), there was a significant reduced mean breathing frequency from T0 (16.60±2.30 breaths per minute) to T1 (15.09±2.27 breaths per minute), following the supervised HRVB training (*p*<.001), but not in the sham group (*p*>.05). None of the other physiological parameters showed significant within-group differences between T0 and T1 (*p*>.05). Within-group differences between T1 and T2 were only present in the Sham+HT group with an increased mean heart rate and reduced mean cardiac vagal modulation (based on logHF-HRV), following the home-based HRVB training (*p*<.05). Regarding the results from the questionnaires between T0 and T1, the HRVB group presented significantly lower self-perceived stress during the Stroop task (5.46±2.25 to 4.38±2.36) and lower self-perceived stress levels according to the PSS (24.08±6.52 to 20.50±6.41) and the stress subscale of the DASS-21 (10.67±4.59 to 8.46±4.72; all *p*<.05). None of the within-group differences on the questionnaires in the sham group were significant (all, *p*>.05). Within-group differences between T1 and T2 were present for the SDQ self-report in the Sham+HT group with a significant decrease from T0 to T2 (*p*<.05). Regarding the SRS total t-score, all groups (except the HRVB+NT group) showed decreased scores at T2 (*p*<.05).

**Table 2.** Physiological and cortisol data reported separately for each assessment session (T0, T1, T2).

|  | Group | T0 | T1 | T0-T1 |  |  | Group | T1 | T2 | T1-T2 |  |  |
| --- | --- | --- | --- | --- | --- | --- | --- | --- | --- | --- | --- | --- |
|  |  | Mean (SD) | Mean (SD) | T(df)/Z | p | d/r |  | Mean (SD) | Mean (SD) | T(df)/Z | p | d<sup>c</sup>/r |
| RMSSD (ms) | Sham | 34.98 (13.36) | 35.41 (16.90) | -0.168(19) | .869 | 0.036 | Sham+NT | 29.84 (6.47) | 29.03 (8.20)<sup>a</sup> | -0.533 | .594 | 0.178 |
|  |  |  |  |  |  |  | Sham+HT | 40.99 (22.18) | 33.26 (20.69) | 2.012(9) | .075 | 0.581 |
|  | HRVB | 44.64 (26.38) | 36.84 (17.16) | -1.600 | .110 | 0.327 | HRVB+NT | 31.13 (18.78) | 40.35 (29.98) | -1.274 | .203 | 0.403 |
|  |  |  |  |  |  |  | HRVB+HT | 40.92 (15.29) | 38.90 (14.19) | 0.471(13) | .646 | 0.118 |
| LogHF-HRV | Sham | 6.28 (0.70) | 6.16 (0.87) | 0.751(19) | .462 | 0.168 | Sham+NT | 5.98 (0.52) | 5.90 (0.68)<sup>a</sup> | -0.652 | .515 | 0.217 |
|  |  |  |  |  |  |  | Sham+HT | 6.35 (1.12) | 5.92 (1.10) | 2.308(9) | **.046\*** | 0.667 |
|  | HRVB | 6.56 (1.12) | 6.28 (0.90) | -1.229 | .219 | 0.251 | HRVB+NT | 5.88 (1.09) | 6.30 (1.15) | -1.677(9) | .128 | 0.485 |
|  |  |  |  |  |  |  | HRVB+HT | 6.57 (0.62) | 6.53 (0.47) | 0.242(13) | .812 | 0.061 |
| HR (bpm) | Sham | 85.31 (7.58) | 84.78 (9.28) | 0.237(19) | .815 | 0.053 | Sham+NT | 86.62 (7.79) | 86.04 (8.32)<sup>a</sup> | -0.296 | .767 | 0.099 |
|  |  |  |  |  |  |  | Sham+HT | 82.95 (10.66) | 86.87 (9.68) | -2.445(9) | **.037\*** | 0.707 |
|  | HRVB | 85.72 (11.19) | 89.52 (12.80) | -1.778(23) | .089 | 0.363 | HRVB+NT | 91.74 (15.76) | 88.78 (12.86) | 0.826(9) | .430 | 0.239 |
|  |  |  |  |  |  |  | HRVB+HT | 87.93 (10.56) | 87.00 (10.38) | 0.375(13) | .713 | 0.094 |
| Breathing frequency (/min) | Sham | 16.68 (2.25) | 16.13 (2.37) | 1.425(19) | .170 | 0.319 | Sham+NT | 16.72 (2.08) | 16.67 (1.80)<sup>a</sup> | -0.935 | .350 | 0.312 |
|  |  |  |  |  |  |  | Sham+HT | 15.54 (2.59) | 15.24 (2.44) | 0.865(9) | .409 | 0.250 |
|  | HRVB | 16.60 (2.30) | 15.09 (2.27) | 4.151(23) | **<.001\*\*** | 0.847 | HRVB+NT | 15.64 (2.27) | 15.48 (3.20) | 0.289(9) | .779 | 0.084 |
|  |  |  |  |  |  |  | HRVB+HT | 14.70 (2.27) | 15.06 (2.66) | -1.118(13) | .284 | 0.281 |
| Cortisol (µg/dl) | Sham | 0.17 (0.14) | 0.18 (0.15) | -0.735(19) | .472 | 0.164 | Sham+NT | 0.21 (0.18) | 0.17 (0.13)<sup>b</sup> | -1.400 | .161 | 0.495 |
|  |  |  |  |  |  |  | Sham+HT | 0.16 (0.12) | 0.13 (0.07) | -0.561 | .575 | 0.177 |
|  | HRVB | 0.12 (0.08)<sup>a</sup> | 0.11 (0.06)<sup>a</sup> | 0.570(22) | .574 | 0.119 | HRVB+NT | 0.12 (0.06) | 0.11 (0.08) | 0.080(9) | .938 | 0.023 |
|  |  |  |  |  |  |  | HRVB+HT | 0.10 (0.06)<sup>a</sup> | 0.11 (0.07)<sup>a</sup> | -0.697(12) | .499 | 0.181 |

a Missing value (n=1); b Missing value (n=2); c Hedges' correction
Sham (n=20), HRVB (n=24), Sham+NT (n=10), Sham+HT (n=10), HRVB+NT (n=10), HRVB+HT (n=14)
T0-T1 differences are based on two groups (HRVB and sham); T1-T2 differences are based on four groups (Sham+NT, Sham+HT, HRVB+NT and HRVB+HT)
Note: HF-HRV=High-Frequency Heart Rate Variability; HR=Heart Rate; HRVB=Heart-Rate Variability Biofeedback; HT=Home-based Training; NT=No Training; RMSSD=root mean square of successive differences; SD=Standard Deviation
Bold values represent significant results: \* p<.05; \*\* p<.01

**Table 3.**
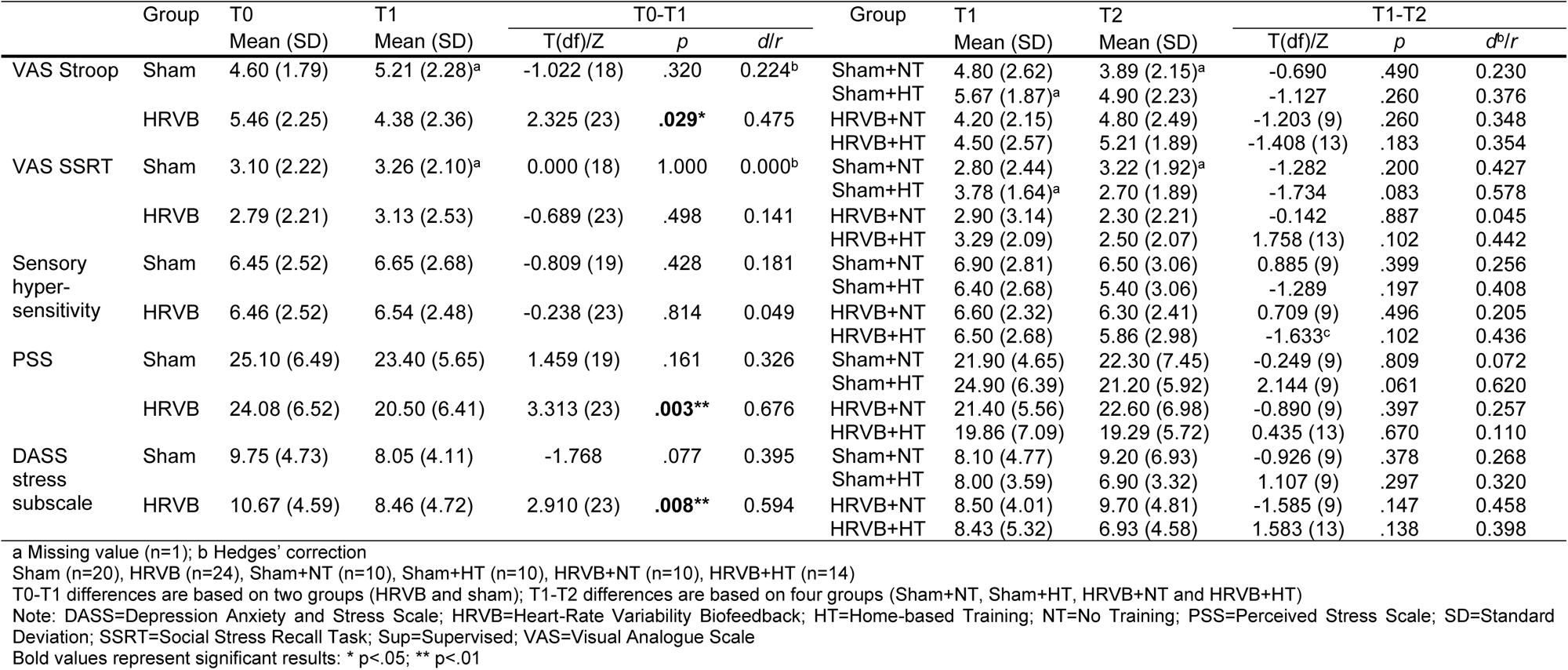
Behavioral data from adolescents with autism reported separately for each assessment session (T0, T1, T2).

**Table 4.**
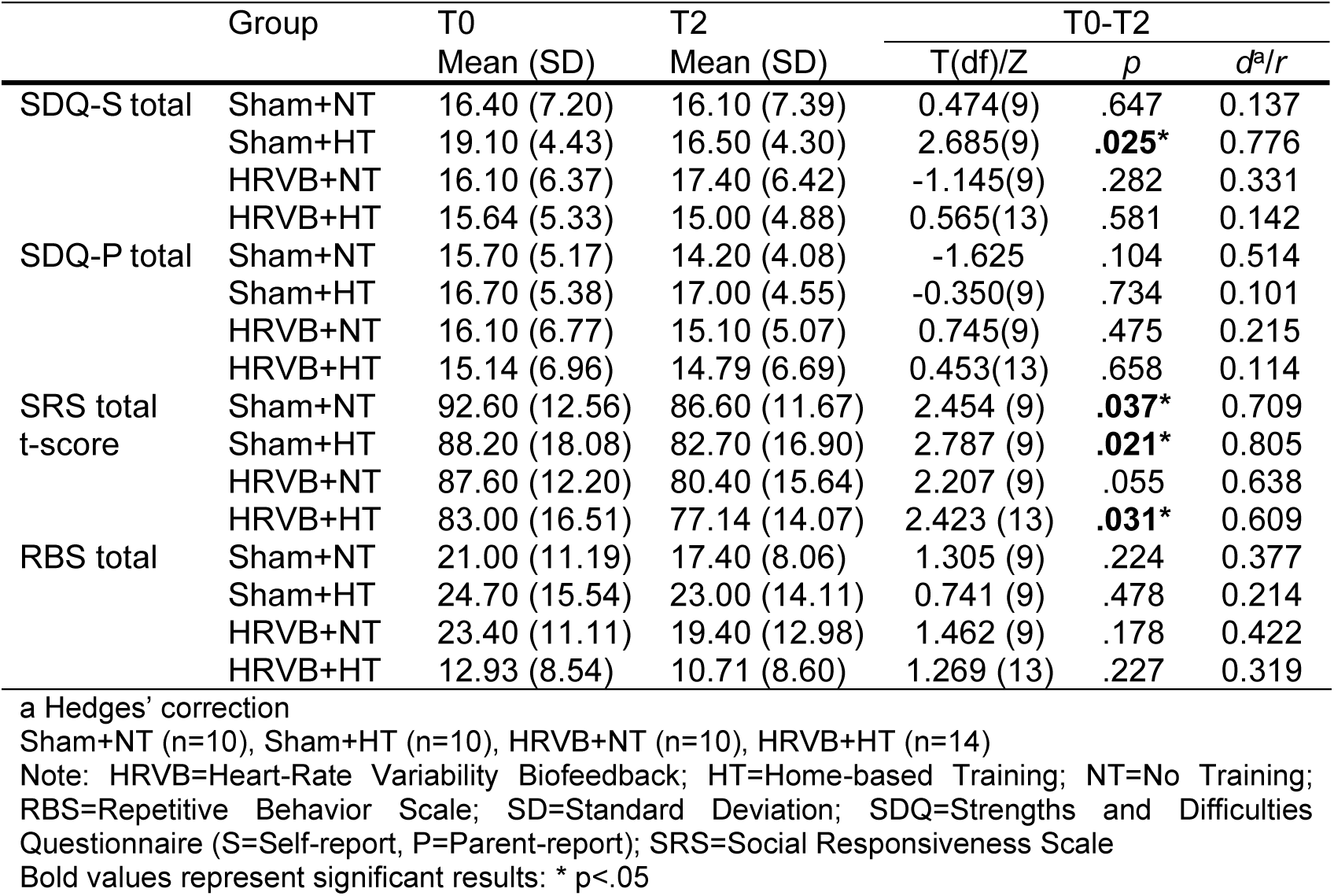
Behavioral data reported separately for assessment sessions T0 and T2.

## Discussion

This pilot RCT study examined whether HRVB training could increase the level of cardiac vagal modulation in adolescents with autism. In addition, the effect on other physiological outcome measures (heart rate and breathing frequency), cortisol levels and outcomes related to psychosocial functioning and self-perceived stress was examined. Further, the feasibility of a home-based, non-supervised HRVB training was determined after the supervised HRVB training. The study findings partially support the first hypothesis, stating that HRVB would result in an increase in cardiac vagal modulation given that an increase in RMSSD was indeed present in the HRVB group with a small effect size in contrast to the sham group. However, this increase was only observed as a late-emerging effect at the T2 follow-up session, five weeks after finalizing the supervised HRVB training period. Regarding the second hypothesis, only heart rate and cortisol levels changed significantly, with a small effect size, immediately following the supervised HRVB training (increase and decrease, respectively), but none of these effects remained significant after five weeks (T2). None of the clinical-behavioral outcome measures on psychosocial functioning and self-perceived stress revealed a significant change following the supervised HRVB training in contrast to the sham group. The last hypothesis stated that a home-based, non-supervised HRVB training would be feasible in adolescents with ASD. The adolescents in this study were able to perform this home-based training, but the compliance rate during this training period dropped significantly. In addition, although most of the outcome measures did not change depending on this additional home-based training, adolescents in the home-based training group did report significantly lower symptoms of stress with a medium effect size (as assessed by the stress scale of the DASS) at T2, regardless of the training that was performed in advance (HRVB or sham).

A recent meta-analysis of Laborde et al. (2022) has focused on the influence of HRVB on the parasympathetic nervous system. They demonstrated that increased vagally mediated HRV, expressed as RMSSD, was present after multiple sessions of HRVB training (or slow-paced breathing). Although the average increase in RMSSD was small after multiple HRVB sessions, the authors suggested that functional changes may indeed occur in vagal nerve efferents (leading to increased HRV). This could have been caused by the frequent stimulation of the vagal afferents during HRVB, thereby confirming the contributing role of the vagus nerve in HRVB (Lehrer & Gevirtz, 2014). The findings of this study are partially in accordance with the ones from Laborde et al. (2022) given the significant increase in cardiac vagal modulation following the supervised HRVB training. However, this effect emerged later since it was only present at five weeks after the training period (T2). As far as the authors know, only two previous studies examined the feasibility and efficacy of HRVB in children and adolescents with autism (Coulter et al., 2022; Goodman et al., 2018). The results of the current study replicate the ones from Goodman et al. (2018) in not finding immediate effects on cardiac vagal modulation following HRVB training. Nevertheless, they did not include a follow-up session, thereby hampering the verification of the proposed hypothesis regarding a late increase of cardiac vagal modulation in adolescents with autism. With regard to other populations, Laborde et al. (2022) reported that 45% of the studies concerning HRVB did not report the influence on cardiac vagal modulation (RMSSD). However, out of the 22 studies that did, three were relevant with respect to the current study as they did not reveal significant changes in RMSSD following multiple sessions of HRVB (Raaijmakers et al., 2013; Winstead, 2019) or breathing at a frequency that maximized HRV amplitude (Tatschl et al., 2020). In the study of Winstead (2019), six weeks of HRVB training (weekly guided sessions and daily home-practice) were performed by veterans (54±11 years). No differences in cardiac vagal modulation were reported at 8-, 12- and 16-weeks post-baseline. Participants in the study of Raaijmakers et al. (2013) were healthy male volunteers (19-27 years) who were asked to engage in a biofeedback game during 7 sessions, spread across 16 days. During this game, both skin conductance and HRV were used as control variables while no home-practice was performed. The authors suggested that HRVB might lead to an overall increase in RMSSD if the feedback on HRV would be used in daily living situations instead of only using it as a control variable during games as in their study. Finally, the study of Tatschl et al. (2020) used a similar training dose as the current study in adults (48.7±9.4 years) who were admitted at an inpatient psychiatric rehabilitation clinic. The authors suggested that the advanced age in their study sample and the density of pharmacological interventions (f.i. antidepressants and antipsychotics) contributed to the lack of change in RMSSD since both factors are known to negatively influence changes in HRV (Alvares et al., 2016; Lehrer et al., 2006; Linder et al., 2014). Given the young sample in this study, the argument of the potential influence of advanced age was not considered. However, the use of psychotropic medication (such as antipsychotics, methylphenidate, and antidepressants) may have moderated the efficacy of the HRVB training since 50% of the adolescents in the HRVB group reported this type of pharmacological intervention. The current study sample was deemed too small to perform moderation analysis on the possible role of psychotropics.

As far as the authors know, this is the first study to suggest a late-emerging effect on cardiac vagal modulation following five weeks of HRVB training. A possible explanation for this later increase might be that adolescents who received the HRVB training implicitly continued to use the learned breathing technique in daily life between T1 and T2, also outside the explicit training context. Although transfer of practice to daily life constitutes an important goal of biofeedback training in general, this study did not include any measurement to verify the extent of transfer. It should be noted that while half of the HRVB group was instructed to proceed with the home-based HRVB training (without supervision), thereby prolonging their explicit training period, this did not result in an additional effect on the level of cardiac vagal modulation at T2. Given that several questions remain regarding the exact working mechanisms of HRVB and the most efficient training protocol (Lehrer, 2022; Lehrer et al., 2020; Pizzoli et al., 2021; Sevoz-Couche & Laborde, 2022), more information on these topics is needed to further explain the late-emerging effects found in this study.

Contrary to the late influence on cardiac vagal modulation, an immediate effect was found on heart rate and cortisol levels in adolescents with autism following HRVB training. While no immediate effect on cardiac vagal modulation was present, the current study showed an increased mean heart rate at T1 in the HRVB group in comparison to the sham group. This was somewhat surprising since the hypothesized increase of cardiac vagal modulation would intuitively result in a lower heart rate. A possible explanation may be that the slow breathing frequency at which the adolescents in the HRVB group had to practice was stress inducing. This may have increased the activity of the sympathetic nervous system in the HRVB group at T1, resulting in an increased heart rate. Exploratory within-group analyses revealed a similar pattern of increased heart rate at T2 for the adolescents who received home-based HRVB after sham training, while this was not present in those who continued HRVB training after supervised training. The inclusion of physiological data on sympathetic nervous system activity in future research is needed to explore this hypothesis of increased sympathetic activity. Five weeks after the HRVB training (T2), the increased heart rate in the HRVB group was no longer apparent, probably due to the increased cardiac vagal modulation with respect to the sham group. Furthermore, the immediate decrease of mean salivary cortisol after HRVB training in comparison to the sham training suggests that HRVB may indeed exert an influence on the activity of the HPA axis. This is similar to what has been found in other studies in adult samples (Bouchard et al., 2012; Brinkmann et al., 2020). This effect, however, was not related to an increase in cardiac vagal modulation, as was hypothesized in the current study. Moreover, at T2, when the effect on cardiac vagal modulation was significant, the difference in cortisol levels between the two groups was only borderline significant. In sum, the current study results suggest that HRVB has a distinct impact on the autonomic nervous system and the HPA axis in adolescents with autism.

The mixed model analyses did not reveal a change in the adolescents’ breathing frequency following HRVB training, despite the significant decrease in breathing frequency for the HRVB group at T1 demonstrated by the exploratory within-group analyses. One explanation may be that the sample size was not large enough; another could be that the decrease in breathing frequency during the HRVB training was not generalized after the training sessions. Of note, a decrease in breathing frequency was only expected for the resting periods, not during the stress-provoking tasks given their verbal character.

The interest in HRVB for various clinical and non-clinical populations across the lifespan has increased exponentially, given the positive effects on physical symptoms, such as asthma and hypertension, and on mental health symptoms such as depression, anxiety and stress (Blase et al., 2021; Dormal et al., 2021; Fernández-Alvarez et al., 2022; Lehrer et al., 2020; Pizzoli et al., 2021; Tatschl et al., 2020; Yetwin et al., 2022). Porges’ Polyvagal Theory (Porges, 2011) and the Neurovisceral Integration Theory (Thayer & Lane, 2000) are often cited in support of these research findings. The Polyvagal Theory proposes that lower levels of cardiac vagal modulation are associated with difficulties in social behavior. The Neurovisceral Integration Theory builds upon the latter theory by describing that cardiac vagal modulation is associated with self-regulatory behavior due to the bidirectional connection between the heart and several related brain regions. Lower levels of cardiac vagal modulation have been found in adolescents with autism in comparison to typically developing peers (Cheng et al., 2020; Makris et al., 2022; Thoen et al., 2023). Thus, it was hypothesized that HRVB would upregulate the level of cardiac vagal modulation in adolescents with autism and would, additionally, result in a decrease of the severity of associated psychosocial and behavioral problems. Of note, the complementary character of HRVB to other well-established and effective psychological treatment modalities has been suggested in the meta-analyses of Lehrer et al. (2020). Therefore, HRVB was considered as a possible support strategy for adolescents with autism to facilitate stress- and self-regulation. However, non-significant changes on psychosocial and behavioral outcome measures were revealed following the HRVB training. This contrasts with the findings of both Goodman et al. (2018) and Coulter et al. (2022), who did report beneficial effects on social behavior, emotion regulation and self-reported anxiety in children and adolescents with autism. It has been suggested that behavioral changes may only become apparent when training duration is increased (Sevoz-Couche & Laborde, 2022). The findings in this study partially confirm this as less stress symptoms, assessed with a number of items of the DASS, were indeed present after the second round of HRVB training (cf. home-based HRVB) but not after the first round (cf. supervised HRVB). It has also been hypothesized by Deschodt-Arsac et al. (2018) that the time course in physiological and behavioral changes resulting from HRVB may be different in a way that physiological changes may precede any changes in behavior. The current study results do not support this hypothesis since the increase in cardiac vagal modulation and the decrease in symptoms of stress were present during the same assessment. In addition, the increase in cardiac vagal modulation was the result of the supervised HRVB training whereas the decrease in stress symptoms was caused by the home-based, non-supervised HRVB training. In line with this, there are studies in which changes in behavior or clinical symptoms were present regardless of any physiological changes (Lehrer, 2022). It is important to mention that half of the adolescents received the sham training prior to the home-based HRVB training. This implies that the effect on stress symptoms in this study cannot be explained entirely by the increased training load of HRVB as not all adolescents received ten weeks of HRVB. It could be argued that the sham training in this study was a type of slow-paced breathing given the slower breathing frequency of the adolescents during the training (10.90±1.77 breaths per minute) with respect to their rather high breathing frequency during the baseline measurement at T0 (17.70±3.31 breaths per minute). Therefore, it could be argued that adolescents in the sham group may have benefited from this training as well. However, the exploratory within-group analyses did not reveal any improvements in self-reported stress immediately after the sham training whereas these effects were present for the HRVB group. None of these immediate effects reached statistical significance in the mixed models (primary analyses), probably due to the small sample size of the groups. Furthermore, no changes were apparent in outcome measures reflecting the impact of autism characteristics or behavioral symptoms in the current study. More research on how HRVB exerts its effects is warranted to explain these inconsistent and seemingly contradictory effects. Finally, at T1, a significant decrease with a medium effect size in sensory hypersensitivity was present in both groups (HRVB and sham training). Since this outcome measure did not assess sensory hypersensitivity across different sensory modes, it would be interesting to explore this valuable effect more in detail in future research.

### Strengths and limitations

This pilot RCT has several strengths such as the inclusion of both boys and girls with autism; a rigorously described assessment protocol that includes the assessment of both cardiac vagal modulation and salivary cortisol levels as well as parent and self-reported outcome measures and the combination of a supervised and home-based HRVB training period. However, some limitations need to be acknowledged as well. One limitation is the small study sample, proper to a pilot study. Replication in a larger RCT is necessary. Furthermore, caution is warranted regarding the interpretation of the exploratory within-group differences since no main effect of group has been found in the mixed-effect models (primary analyses). However, these exploratory analyses were included for completeness and to determine whether possible trends were present within the groups that could not be detected in the primary analyses due to the small group sizes. Possible moderating effects such as sex, presence of co-occurring disorders or psychosocial symptoms, use of psychotropic medication and severity of autism characteristics were not included in the analyses since the study sample was deemed too small to perform moderation analyses. Of note, a previous study using a subgroup of the current sample of adolescents with autism revealed a non-significant effect of psychotropic medication on the level of cardiac vagal modulation during the stress-provoking protocol (Thoen et al., 2023). However, more information regarding possible moderating factors is valuable to further define who would benefit most from HRVB and to determine whether personalized HRVB protocols are needed to maximize training-related benefits (Lehrer, 2022; Pizzoli et al., 2021). Finally, this study only included adolescents with autism without intellectual disability (ID) despite the preliminary evidence for the feasibility of slow-paced breathing in adolescents with ID (15-19 years), as confirmed in a pilot study of Laborde et al. (2017). Of note, their training protocol was based on breathing at a frequency of six breaths per minute, which is close to the mean resonance frequency found in adults, thereby relying on a similar working mechanism as HRVB. Short-term increases of cardiac vagal modulation during a cognitive task were present in their slow-paced breathing group in contrast to the comparison group that listened to an audiobook. However, they recommended more research given their small sample size, the preponderance of males in their study population and the exploration of short-term effects only (Laborde et al., 2017). Given the large co-occurrence of ID in individuals with autism, it would be interesting to further expand research on HRVB efficacy in this population.

### Directions for future research

Suggestions for future research are in place to further examine the efficacy of HRVB in adolescents with autism and on HRVB in general.

First, regarding the study design, it might be valuable to include a follow-up assessment, such as in this study, to determine whether late influences on physiological and/or behavioral parameters are present (Goessl et al., 2017; Lehrer et al., 2020). In addition, a longer training period has been suggested to result in better clinical outcomes (Blase et al., 2021), but currently there is no consensus regarding the best training protocol to obtain long-lasting effects after HRVB training (Lehrer, 2022; Pizzoli et al., 2021).

Second, given the significant decrease in compliance rate during the home-based HRVB training period in this study, it might be valuable to include virtual follow-up sessions to motivate adolescents with autism in continuing with the training. Furthermore, it has been suggested to use different modes of feedback during HRVB (e.g., tactile, auditory, and visual feedback) in addition to a personalized interface (Fernández-Alvarez et al., 2022; Morales et al., 2022). In a recent study in children with autism (6-10 years), an adaptive biofeedback model was proposed which integrates physiological data, therapy adherence and environmental data during breathing exercises in order to provide personalized feedback (Morales et al., 2022). Despite the need for further development of this adaptive model, it might be valuable for adolescents with autism as well. Although many of the adolescents in this study did not report frustrations upon using the breathing pacer applications, some of them did mention that using more engaging interfaces could have increased their therapy motivation and compliance.

Third, collecting objective data regarding training compliance would be beneficial in future research. For instance, using devices that capture data during home-training sessions could provide researchers with accurate information regarding training duration and timing. In this study, a similar device (Polar heart rate sensor) and application (Elite HRV) were originally described to use during the home-based training period. However, technical problems and practical issues (e.g., difficult to apply the heart rate sensor across settings) prevented multiple adolescents from using them. Therefore, they continued using the previously installed breathing pacer applications (from the supervised training period) which hampered the objective data acquisition. This contrasts with the findings of Coulter et al. (2022), who reported good usability of two HRVB devices in young individuals with autism. However, these devices were based on infrared photoplethysmography (PPG) sensors that were placed around one’s finger or earlobe, thus are easier to use as opposed to the chest belt used in this study. Additionally, qualitative interviews regarding the usability of HRVB devices and applications as well as the tolerance of the HRVB training in adolescents with autism would complement the objective information (Yu et al., 2018).

Fourth, including an outcome measure that targets immediate session-related effects on self-perceived stress could provide valuable information as well. The reasoning behind this is based on the hypothesized induced sense of relaxation following HRVB training since the vagus nerve is stimulated (Lehrer et al., 2020).

Fifth, this study included a sham training group to control for the possible placebo effect of breathing awareness or distraction during HRVB (Sevoz-Couche & Laborde, 2022). However, HRVB might be beneficial in adolescents with autism in comparison to other stress-management techniques or methods (e.g., mindfulness). Therefore, it would be valuable to explore this by including additional control groups based on these techniques.

Finally, it has been recommended to further elucidate the possible working mechanisms of HRVB (Goessl et al., 2017; Lehrer, 2022; Lehrer et al., 2020; Sevoz-Couche & Laborde, 2022). The inclusion of outcome measures that reflect the functioning of both branches of the autonomic nervous system as well as measures reflecting the functioning of the HPA axis might provide additional information regarding physiological changes caused by HRVB. Recent studies also indicated that HRVB may enhance resting state oscillatory blood flow and brain connectivity in cortical areas related to emotion regulation (Mather & Thayer, 2018; Nashiro et al., 2023; Schumann et al., 2021). This might explain the various positive behavioral changes following HRVB (Lehrer et al., 2020). However, future research should incorporate outcome measures that can capture brain-related changes to further elucidate and confirm this hypothesized working mechanism.

## Conclusion

The findings from this pilot RCT suggest that HRVB is feasible and effective in adolescents with autism, demonstrated by late-emerging increases in cardiac vagal modulation and decreases in self-reported stress symptoms. Future researchers are encouraged to include larger samples to verify these results as well as including moderation analyses to explore which factors may influence treatment efficacy. Finally, as has been proposed previously, including additional outcome measures to further elucidate the possible working mechanisms of HRVB is recommended.

### CRediT author contribution statement

**Anoushka Thoen:** Conceptualization, Methodology, Formal Analysis, Investigation, Data Curation, Writing - original draft, Visualization, Project administration, Funding acquisition. **Kaat Alaerts:** Conceptualization, Methodology, Software, Formal Analysis, Resources, Writing - review & editing. **Jellina Prinsen:** Software, Writing - review & editing. **Jean Steyaert:** Conceptualization, Resources, Writing - review & editing. **Tine Van Damme:** Conceptualization, Methodology, Resources, Writing - review & editing, Supervision.

### Data availability

Data will be made available on request.

## Data Availability

All data produced in the present study are available upon reasonable request to the authors

## Acknowledgements

This work is supported by the Marguerite-Marie Delacroix foundation with grant number RVC/B-472, awarded to AT. The funder did not have any role in any part of this study. The authors wish to thank the members of the lab of Professor Verhaeghe (UZ Leuven, Belgium) for performing the cortisol analyses. Miss Angeliki Karaiskou and Miss Naishi Feng are acknowledged for their contribution to the extraction and preprocessing of the respiration data. Miss Sophie Pleysier is acknowledged for her contribution in digitalizing the research notes. The sponsor of this study is KU Leuven (Oude Markt 13, 3000 Leuven, Belgium) and will have no role in any part of this study.

## Supplements

**Supplement 1.**
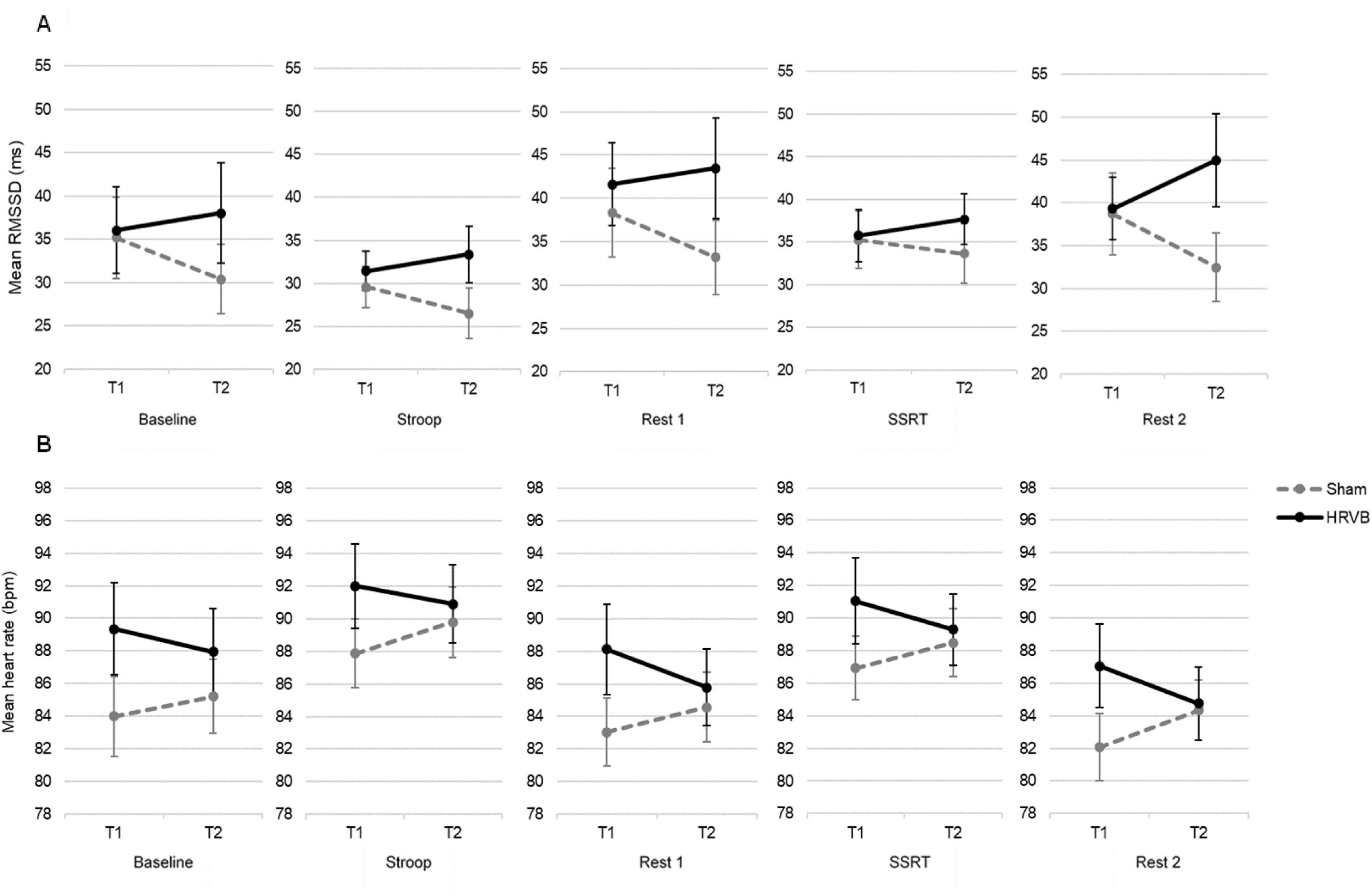
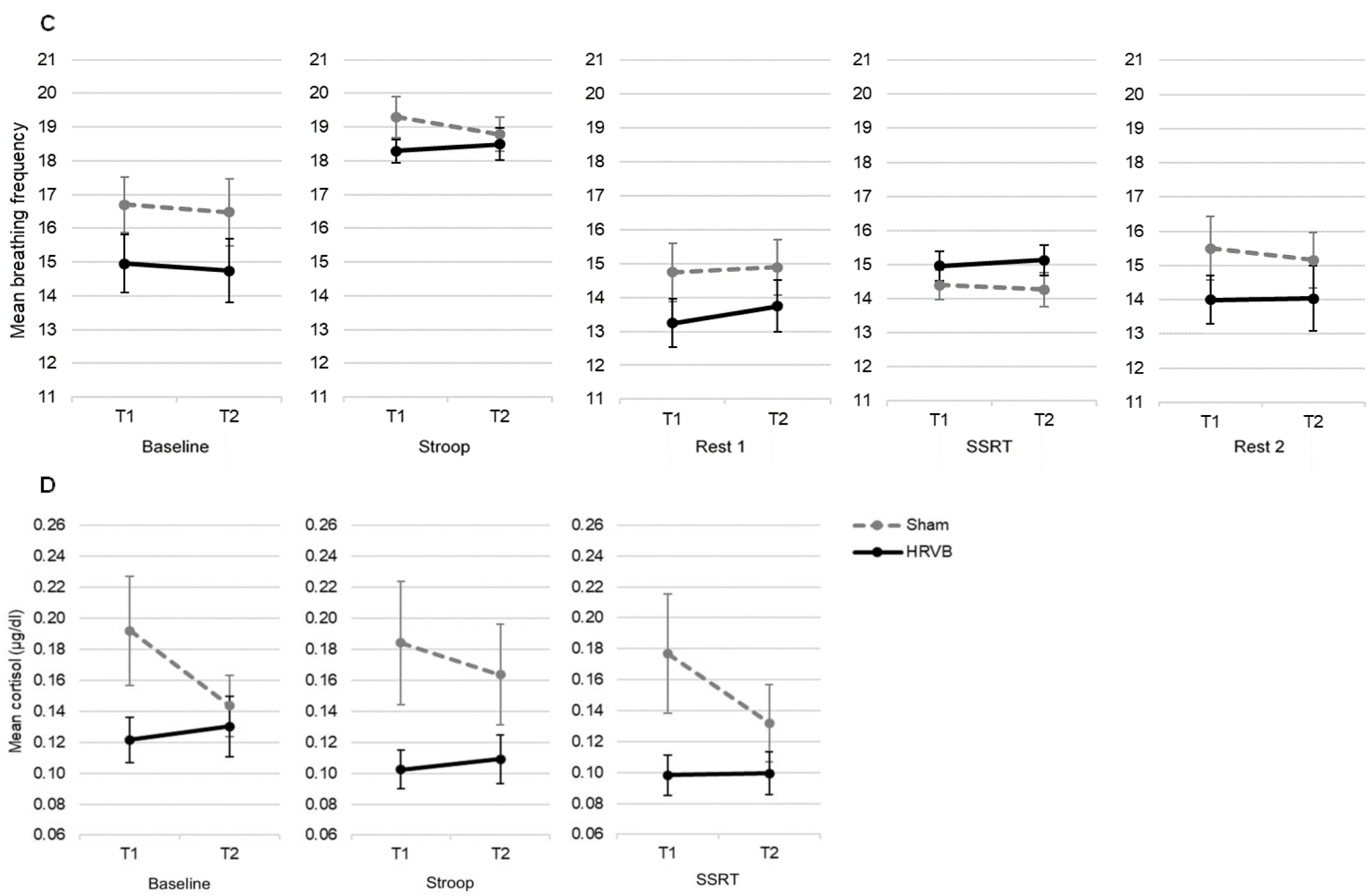
Means are visualized per condition, separately for the HRVB and sham group at assessment sessions T1 and T2. Panel A: Effect on cardiac vagal modulation, assessed using RMSSD. Panel B: Effect on heart rate. Panel C: Effect on breathing frequency. Panel D: Effect on cortisol level. Note: bpm=beats per minute; HRVB=Heart Rate Variability Biofeedback; RMSSD=Root Mean Square of Successive Differences.

**Supplement 2.**
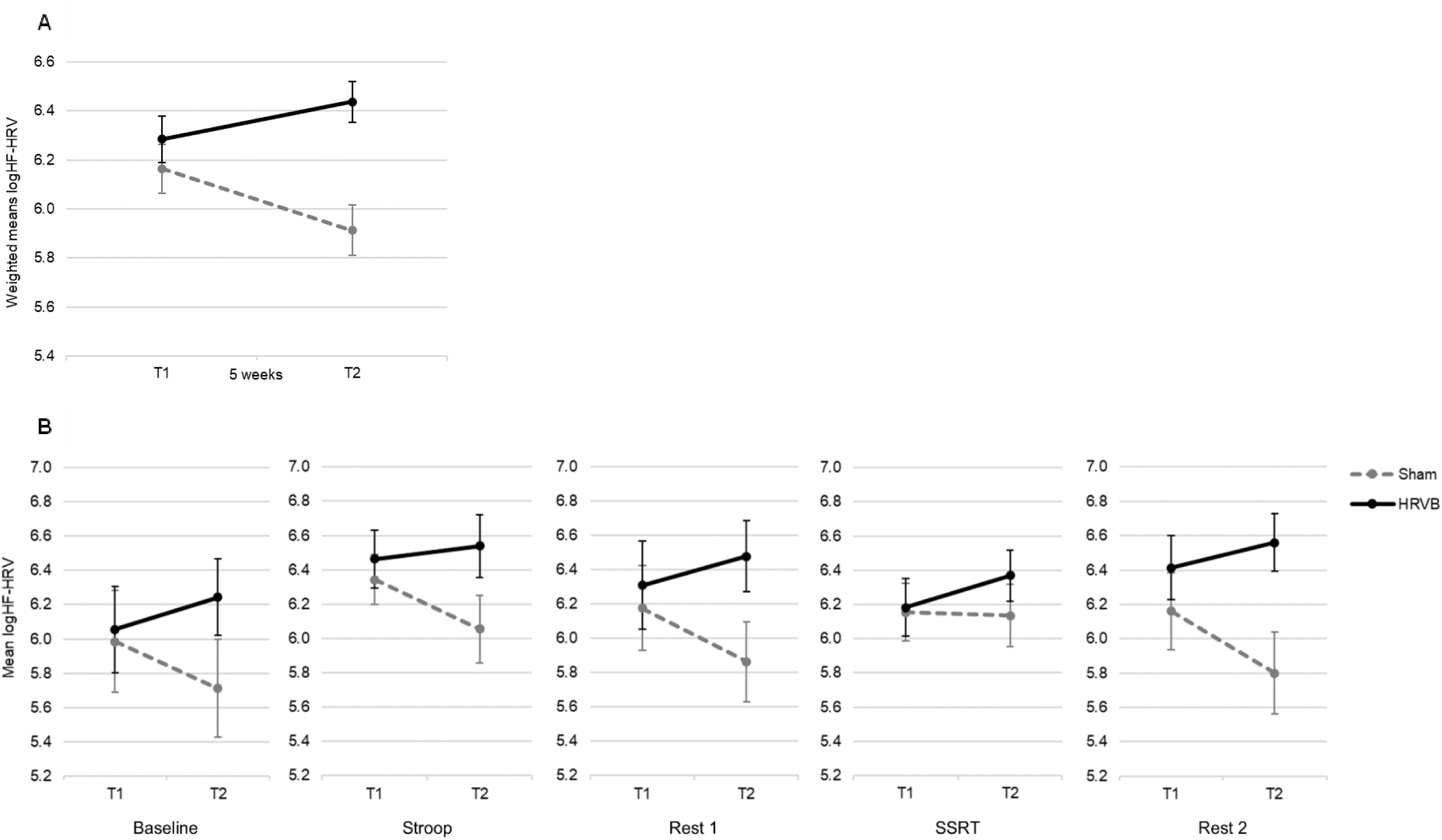
Means of logarithm-transformed HF-HRV are visualized separately for the HRVB and sham group at assessment sessions T1 and T2. Panel A: Weighted means. Panel B: Means per condition. Note: HF-HRV=High-Frequency Heart Rate Variability; HRVB=Heart Rate Variability Biofeedback.

**Supplement 3.**
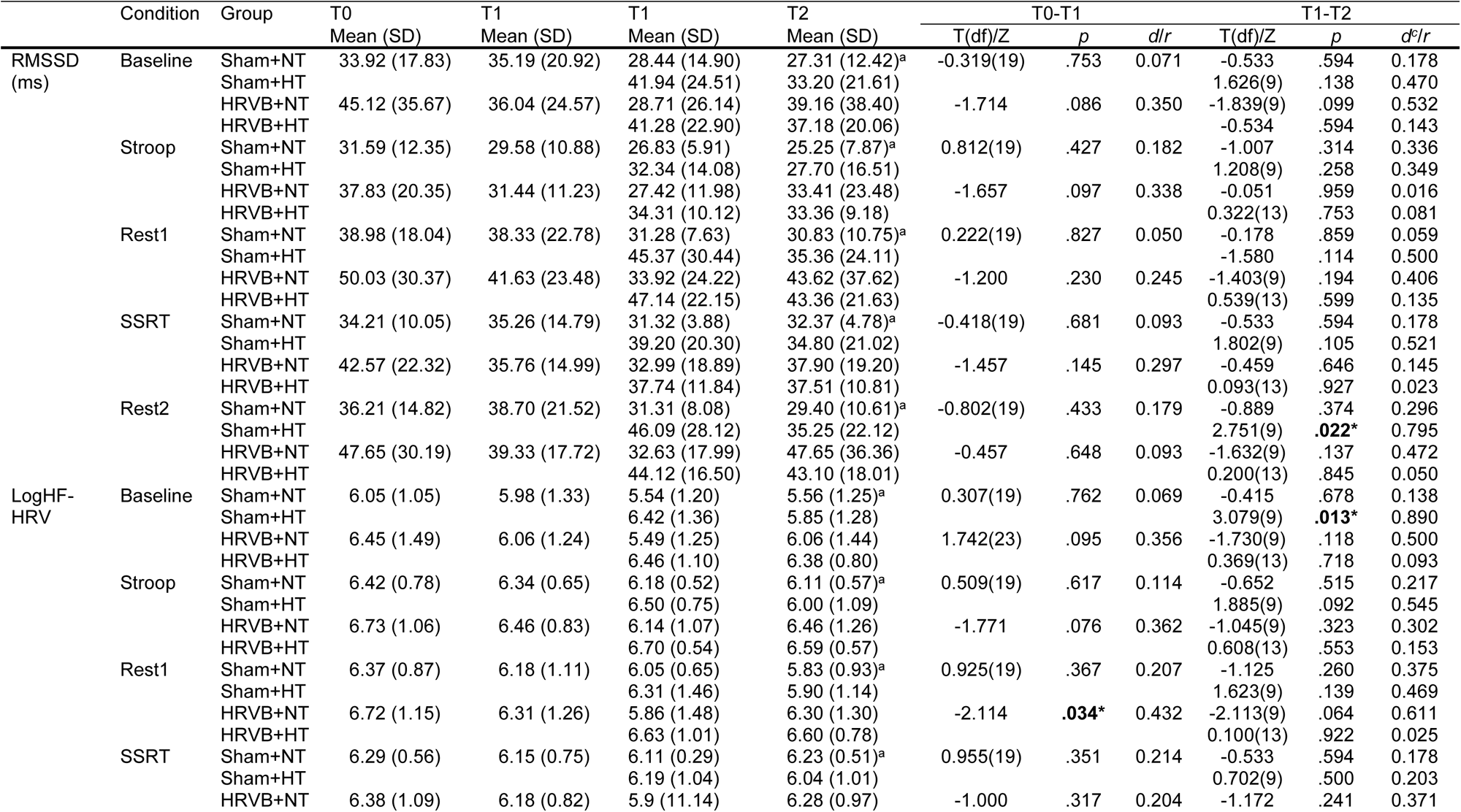
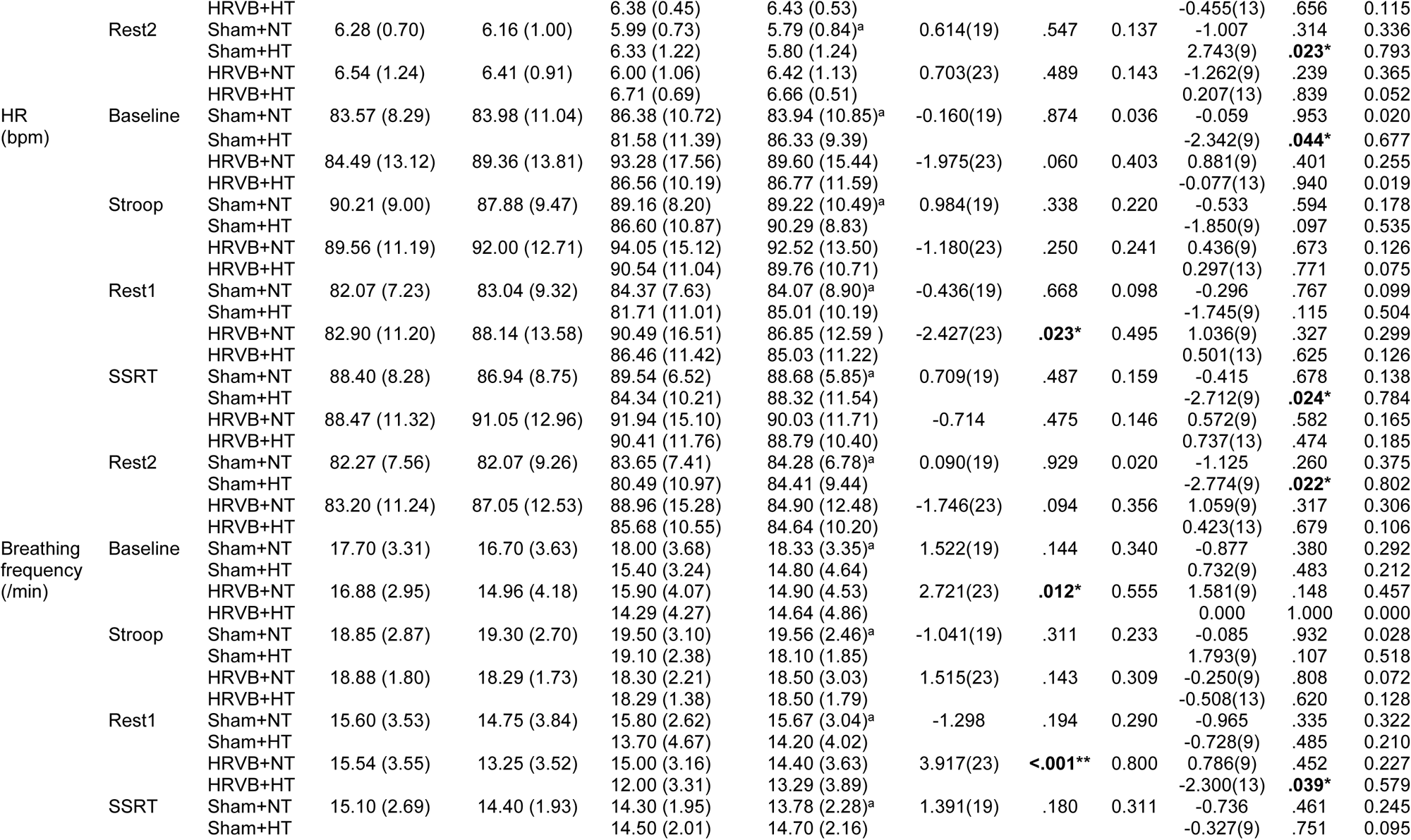
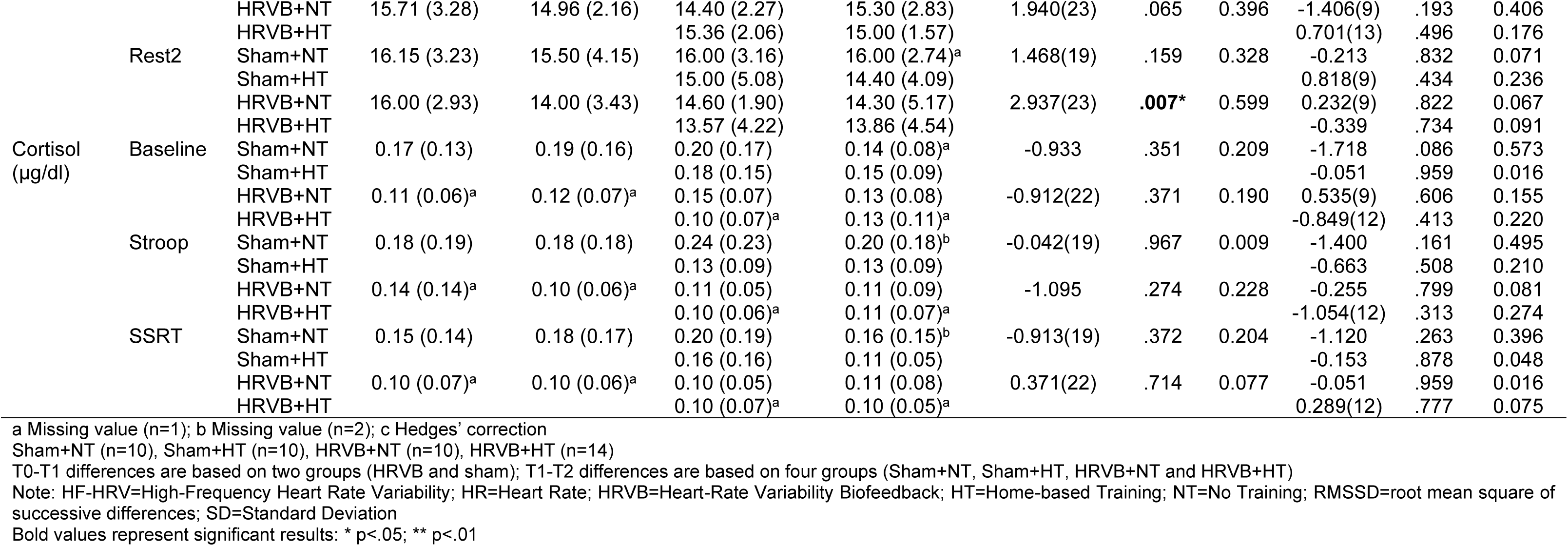
Physiological and cortisol data reported separately per condition for each assessment session (T0, T1, T2).

## Footnotes

1 The authors recognize that language preferences are both personal and culturally bound. In this manuscript, person-first language is used as this seems to be preferred by the majority of Dutch-speaking people with autism (Buijsman et al. 2023). The current ICD-11/DSM-5 diagnostic terminology of “Autism Spectrum Disorder” (abbreviated as “ASD”) is used when referring to the diagnostic criteria or process.

